# Volunteers of Third Sector Organizations in supporting older adults in the transition from hospital to home: A comparative case study

**DOI:** 10.1101/2023.06.05.23290992

**Authors:** MLA Nelson, H. Singh, M. Saragosa

**Affiliations:** Science of Care Institute; Lunenfeld-Tanenbaum Research Institute, Sinai Health; Department of Occupational Science & Occupational Therapy, Temerty Faculty of Medicine, University of Toronto, Toronto, Ontario, Canada; Institute of Health Policy, Management, and Evaluation, Dalla Lana School of Public Health, University of Toronto, Toronto, Ontario, Canada

**Keywords:** Transitions, third sector, case study, intermediate care, hospital discharge, person centred care

## Abstract

**Introduction:** With increasing attention to models of transitional support delivered through multisectoral approaches, third sector organizations have supported community reintegration and independent living post hospitalization. This study aimed to identify core elements of these programs, facilitators and barriers to service implementation, and to understand the perspectives of providers and recipients.

**Methods and Analysis:** A comparative case study was conducted, collecting data from two ‘Home from Hospital’ programs in the UK, and two transition support programs in Canada. An inductive thematic analysis generated rich descriptions of each program, and comparative analytical activities generated insights across the cases.

**Results:** Programs provided a range of personalized support for older adults and address many post-discharge needs, including wellbeing assessments, support for instrumental activities of daily living, psychosocial support, and other individualized services directed by the needs and preferences of the service user. Results suggest that these types of programs can act as a ‘safety net’ and promote independent living. Skilled volunteers can positively impact older adults’ experience returning home.

**Conclusions:** When the programs under study are considered in tandem with existing evidence, it facilitates a discussion of how TSO services could be made available more widely to support older adults in their transition experiences.

## Background

Many people transitioning home after hospital discharge, particularly older adults, continue to experience significant unmet health care demands as they readjust to life at home (1-4). Unresolved somatic symptoms, difficulties in daily management and processing of information, and an insufficient social support network contribute to challenging transitions home (5). Recognizing that well-facilitated and supported transitions can decrease adverse events and re-hospitalization (6-9) and that health, practical, and social supports are needed to manage the transition home from the hospital, various health service models have been developed to address these needs (10, 11). There has been increasing attention to models of transitional aftercare delivered through multisectoral approaches (9), defined as the deliberate collaboration among various stakeholder groups, and sectors, including public (e.g., hospitals and health authorities), private (e.g., for-profit organizations), and in some countries, third (e.g., voluntary organizations) sectors (12, 13). In some jurisdictions, third sector organizations (TSOs)– have stepped in as providers of community reintegration and independent living post hospitalization services (14).

The term ‘third sector’ describes organizations that belong neither to the public (e.g., the state) nor the private (e.g., for-profit private organizations) sectors. Coined to describe the UK and North American organizations, TSOs could include non-profits, think tanks, registered charities, and other organizations such as self-help groups, and social enterprises as a sector, distinct from public and private sectors (15). Regardless of jurisdiction, however, the TSOs share five general characteristics (16):

1. Organized: TSOs operate as established institutions.
2. Non-governmental: although TSOs may collaborate with government agencies and receive government funding, they remain independent from the government.
3. Non-profit distributing: TSOs do not operate to generate profits; funds are raised to invest in health, social, environmental or cultural objectives.
4. Self-governing: TSOs operate independently without interference from external authorities.
5. Voluntary: TSOs are driven by the commitment of individuals operating within it and typically involve a meaningful degree of volunteerism.

Collaboration and partnerships between health care organizations and TSOs may reduce demands on health systems, as engaging multiple sectors may be an opportunity to leverage existing knowledge, expertise, and resources to support the shared goal of improving health outcomes (17). Newton noted that third sector organizations are well placed to support people regain independence at home, and often have a track record in providing it: “Formal partnerships between reablement providers and voluntary organizations are likely to be the way forward” (12).

Despite the engagement of TSOs in providing supports research on the delivery of transitional care has predominantly focused on the role of nurses, health disciplines, and other formal health care providers (18, 19), with limited attention given to volunteers at health-based TSOs. Specifically, the role, contributions, and value of TSOs in delivering transitional care services have received less attention and need to be clearly and consistently defined. Research on the engagement of volunteers and partnerships with TSOs to support care transitions and independent living at home is emerging (14).

Recognizing the potential value of TSOs in improving client experience and outcomes, British and North American think tank organizations, such as The Kings Fund, the Institute for HealthCare Improvement, and The Beryl Institute, have advocated for more purposeful engagement of TSOs in the design and delivery of health services (20-22).

This paper synthesizes evidence from four case studies of volunteer-provided transition support programs providing intermediate care services for older people post-hospital discharge. This study aimed to identify the core elements of these programs, the facilitators and barriers to service implementation, and the perspectives of providers and recipients regarding these services.

## Methods

### Design

A comparative case study was conducted with research ethics approval from <blinded for review>.

### Recruitment

An environmental scan and referral sampling identified nine volunteer-provided transitions support programs that met the following criteria. The programs i) were delivered by TSOs, with volunteers as the primary service provider; ii) were focused on returning to independent living post-hospitalization; iii) provided services to an older adult population; and iv) were an established model of service (i.e., not a pilot/demonstration project or a part of a research study). Four TSOs that provided transitional support programs agreed to participate: two ‘Home from Hospital’ programs were in the UK, and two transition support programs, including a peer support program and a navigation and community reintegration support service, were in Canada.

### Data collection

A multi-method data collection strategy was employed. Qualitative data were collected on each program, including perspectives on the value of the program or service by the administration, volunteers, clients, and the successes and challenges they experienced related to the program or service. Interviews and focus groups were conducted with 54 participants (15 program staff, 22 program volunteers, and 17 clients). Interviews were recorded, transcribed, and de-identified. Program documents were also collected by retrieving public information about the programs and administrative documents provided by program staff. Although each program evaluated its services annually, it generally consisted of client and volunteer satisfaction surveys; measures were not standardized and did not record relevant health outcomes consistently. Therefore, these data are not reported here.

### Data analysis

Data analysis and collection occurred concurrently while the researchers created case summaries and wrote analytical memos using the transcripts. An inductive thematic analysis was applied by identifying, analyzing and reporting patterns (themes) within the data (23). As we employed an inductive approach, using an in vivo approaches, we identified codes and categories used for analyses directly from the interview data (24). As a result, we were able to generate a rich and detailed account of the cases (23). Common threads were searched for and identified across the entire data set of the interviews (25). The documents were analyzed through an iterative process of skimming (superficial examination), reading (thorough examination), and interpretation (26). Elements of content analyses were used, a first-pass document review was conducted, and meaningful and relevant passages were identified (26).

## Results

Four de-identified programs were included in this study. The included programs had similar missions and objectives: to provide support and services at different points during a person’s transition between hospital and home. All programs used volunteers as the primary ‘workforce’ to support individuals and had formalized intake mechanisms (e.g., recruitment, screening, interviewing) and training processes for the volunteers. The programs recognized the value of utilizing volunteers as the primary ‘program delivery workforce,’ with the supply of volunteers ranging from 15 to 70 individuals. Programmatic details can be found in Table 1.

**Table 1:** Program characteristics of case studies

| **Program** | ***UK Program 1:***<br>***Home from***<br>***Hospital***<br>***Service*** | ***UK Program 2:***<br>***Home from***<br>***Hospital Service*** | ***Canadian***<br>***Program 1:***<br>***Peer Support***<br>***Service*** | ***Canadian Program***<br>***2:***<br>***Navigation and***<br>***Community***<br>***Reintegration***<br>***Support Service*** |
| --- | --- | --- | --- | --- |
| **Program Purpose** | Help older adults as they transition from the hospital to home and regain independence | Provide older adults transitioning home from hospital services | Provide peer support to people who have experienced a stroke or are caregivers | Support older adults who have experienced a stroke and family caregivers through the transition from hospital to home and community |
| **Staffing Model** | 2 full-time dedicated staff, with approximately 70 volunteers | 3 staff (full and part time), with approximately 50 volunteers | Regionally managed by paid staff, variable number of volunteers from within each local community | 1 contract staff, with approximately 15 volunteers |
| **Program Clientele and Admission Criteria** | People over 55 years of age who live alone, and are unable to live independently in the short term | People over 75 years of age who live alone or with a carer, have limited or no social support, and no social care package | People who have had a stroke or their family caregiver. Clients were primarily older adults (over 65 years of age) | Older adults (60 years of age or older) with stroke who lived in the community prior to hospitalization, caregiver available and interested, and resided in the service catchment area |
| **Services Provided** | 8-week time limited service provided. Needs assessment conducted. Support included: shopping, light housework, aid to attend healthcare appointments, collecting prescriptions, emotional support, and signposting to other services | 8-week time limited service initiated prior to hospital discharge with an assessment of need. Assistance with meal making, dog walking, gardening, shopping, collecting prescriptions, transportation to appointments, seated exercises, and befriending | Individualized emotional support to people with stroke and their family and/or caregivers provided through hospital visits and community support groups | 8-week program had several elements: linking stroke survivors and caregivers with community resources; helping them navigate community services; monitoring the implementation of client service and goal plans; creating a community directory; and providing education |

### Core Program Components

Each included program was purposefully designed to support people in regaining the confidence and skills required to live independently and remain socially engaged in their community. Each program supported older adults (55 or older) transitioning home from the hospital post-discharge. All programs included recruited volunteers from their respective communities.

Three of the four programs were designated as time-limited services, targeting the transition from hospital lasting up to eight weeks (UK Program 1 and 2; Canadian Program 2). The fourth program (Canadian Program 1: Peers Support Program) provided services at discharge from the hospital and when clients were living in their homes, transitioning clients to group-based peer support programs. Each program provided its services within the clients’ homes and communities. Although clients were not expected to travel to an office or community setting to participate in the program, many were accompanied by volunteers on shopping excursions or received transportation to and from medical appointments. Lastly, each intervention was tailored to the goals and needs of the clients to support their return to independence and social participation.

### Perspectives of Program Providers (Staff and Volunteers) and Clients

The program developers, coordinators, volunteers and clients provided valuable insights into their experiences with the program and what contribution these types of programs made to supporting independent living and social engagement in the community. Four broad themes were determined i) the programs are perceived to provide a ‘safety net’ for clients and reduce the burden on friends and families, ii) psychosocial aspects of the service are highly valued, and iii) the service fills a gap between health settings and community, and iv) the programs have the potential to improve key health outcomes.

#### Theme 1: Programs provide a ‘safety net’ for clients and reduce the burden on friends and family

Many clients stated that they preferred not to rely on friends and family for support as they transitioned from hospital to home. Instead, they were more comfortable receiving support through a formalized volunteer service that could help with daily tasks until they could do those activities independently. When we asked a client how she would cope if she did not have a volunteer and what she thought her recovery would look like, she stated:

> I dread to think I would just have been reliant on friends, and you’d just get more and more embarrassed about what they were doing for you because you can’t keep asking. I mean 6 weeks is a long time to have someone that you’ve got to keep saying, “Can you do this for me? Can you do that for me? (Client: UK Program 2: Home from Hospital).

Moreover, receiving services from a volunteer vetted by a trusted community organization instilled a sense of comfort. One client expressed, “I just knew that they would send you someone that you could trust.” The engagement of an established community organization also provided comfort and relief to families and caregivers of clients. She also shared that her daughter felt more at ease knowing that she had help:

> Oh yes, it’s taken a lot off her mind knowing that I am being looked after somewhere along the line. If it’s not the district nurses, I’ve got <organization name> people. You know, somebody coming in to look after the sanitary sort of things. So it’s taken a lot off her mind. She’s got enough problems of her own (Client: UK Program: Home from Hospital).

Although safety and risk reduction were assessed in all the programs, the clients of the UK Home from Hospital programs underwent formal risk assessments, including health and wellbeing assessments home and environment/safety checks. Table 2 summarizes the risks and conditions assessed by the home from hospital services.

**Table 2:** An overview of the assessment of risks and concerns within the UK programs

| UK Program 1<br>(Home from Hospital Service) | UK Program 2<br>(Home from Hospital Service) |
| --- | --- |
| -Assessment checklist includes the client's self reported considerations of their health (emotional and physical), environment (situation and condition), resources (financial, home, care and travel), background and social relations (family/neighbours/friends supporting-separation and bereavement)<br>-Health and safety check sheet includes assessment of outside property, inside property, and other areas as required | -Assessment checklist includes self reported considerations of health and wellbeing, emotional health, social connectedness, practical support<br>-Referral forms and first contact checklists consider and assess health, wellbeing, safety and security. Specifically, staying well at home, feeling safe at home, managing money well, and staying active, social and healthy |

#### Theme 2: Psychosocial support a key program component

Many program volunteers noted that clients often expressed loneliness and were usually anxious about returning home from the hospital. The lack of social support or friends and family living a considerable distance away or unavailable for daily or weekly visits increased their worry. As a result, social support or the concept of ‘befriending’ seemed to be one of the more valuable components of the programs.

> Its’ nice, the volunteer comes around once a week and helps me out with the sweeping up, and other odd things. But the nicest part is the chat we have over a cup of tea. She has time to talk and I always feel better when she has been here (Client: UK Program 1: Home from Hospital Program).

In addition, many of the program coordinators described befriending as a program foundation, building a relationship with the client and encouraging them to develop their confidence to physically move about their homes and engage in instrumental activities of daily living.

Another coordinator that managed peer volunteers explained the importance of psychosocial support for the clients they support, augmenting physical rehabilitation and functional recovery services:

> They can do physical rehab, but that doesn’t mean they’re going to adjust and do well. They might walk but that doesn’t mean they’re doing well. You have to think about that psychosocial piece (Program Coordinator: Canadian Program 2: Navigation and Community Reintegration Support Service).

Participants from the Peer Support Service program in Canada discussed how ‘hope’ is important for clients anxious about their transition home and navigating in their communities. Since volunteers in this program were also individuals who have had a stroke, the connection through lived experience and role modeling could inspire and instill confidence in the client:

> One participant told me that a visit from the peer volunteer had more impact on their hope for the future than the last 6 weeks of rehab because that volunteer gave them hope and inspiration and motivation. You know, that’s incredibly valuable (Program Coordinator: Canadian Program 1: Peer Support Program).

#### Theme 3: ‘Bridging the gap’ between hospital care and independent living

The UK-based services described the programs as filling the gap between the hospital, home and community. Program volunteers discussed how they help ensure that support is re-established when individuals return home. In that sense, they viewed themselves as more than volunteers, they were carers:

> This particular client had just been discharged from hospital. So she’s still a bit confused. So I made a few phone calls to make sure that she had the support re-established. And I think our role essentially is that… we’re carers at the moment. There’s lots of different aspects, and it’s all been fragmented so I am filling the gap. I’m forever trying to liaison on their behalf to GPs and nurses (Volunteer: UK Program 1: Home from Hospital Service).

Community reintegration, resuming social activities, and community engagement were important goals for clients and were emphasized by the program coordinators. A program coordinator noted that peer visitors and community navigators could facilitate the connection to community resources:

> Making contact with someone from the community while they are in hospital, or as soon as possible is important in terms of community reintegration. So the peer visitation program is about having that connection, that friendly face. And what we’ve done is that we’ve developed a process where people get referred to a peer visitation program, and the navigator (Volunteer: Canadian Program 2: Navigation and Community Reintegration Support Service).

In addition to peer support for community connection, the programs included a formal signposting or referral process to other community resources and services based on client needs. As the time-limited program was ending, the program staff would discuss any ongoing needs the client may have and signpost the individuals to other longer-term services in the community.

#### Theme 4: Perceived potential for improved outcomes

Many coordinators of the programs believe that hospital discharge could be expedited, or hospital readmissions could be reduced through these types of transition support programs. Program coordinators considered how various supportive tasks volunteers performed could be linked to improved outcomes.

> So I mean that’s costing the hospitals hundreds of pounds a night to keep them in hospital when they don’t really need to be there. They’re ready to go home. And I’m sure I don’t need to tell you guys that the last thing they want is to stay longer than necessary because every day they’re deteriorating mentally and physically a little more. So you know, if we can help out with discharge support, then we do try to…We try to use our volunteers as much as possible (Coordinator: UK Program 2: Home from Hospital).

While program developers believed that client outcomes could be improved with their provided services, they also noted that these outcomes were not systematically or consistently measured and reported. Demonstrating impact is particularly important to secure funding from health authorities, donors and other sources. One Home from Hospital coordinator noted:

> Reducing social isolation has an impact on readmission to hospital. So does reducing dropped medical appointments and picking up on things that have got interfered with by admission to hospital. And reducing risk of falling… these are things that we try to look at and follow up on. It’s really important that we ask about it and measure it. We try to be as accurate as possible in finding out what readmissions there have been and recording them and working to reduce it whatever way we can (Coordinator: UK Program 1: Home from Hospital).

## Discussion

This study describes four programs delivered by TSOs designed to support older adults transitioning from hospital to home and a return to independent living, in two jurisdictions: Canada and the UK. The programs provided a range of personalized support for older adults through services focused on instrumental activities of daily living and psychosocial supports which address many of the post-discharge needs well documented in the literature (1, 27-29), including wellbeing assessments, support for instrumental activities of daily living, psychosocial support, and individualized services directed by the needs and preferences of the service user. The programs were not commissioned to provide any personal care support (hygiene, medications, etc.) and these activities were out of scope for the service. Although each program evaluated its services, it was generally a process and satisfaction survey; measures were not standardized and did not consistently record relevant health outcomes. However, the program managers, coordinators, volunteers, and clients strongly endorsed the value of these types of programs in helping the return to independence at home. Therefore, these results provide an opportunity to discuss the role of TSOs in transitional care, bridging the gap between hospital and home and/or community and the benefits to older adults, their families and the health care systems.

It is well documented that intersecting health and social issues often complicate the transition process and adversely affect physical recovery, mood, social participation, quality of life and ultimately, the self-management and self-care capacity of patients and caregivers (18, 30-32). The clients of the included programs benefited from a range of instrumental activity supports, such as transportation, grocery shopping, light housekeeping, and appreciated the psychosocial support provided by program volunteers. A key finding of this research was the perceived value of older adults not having rely on family and friends to meet these needs, and the perception that this was a benefit to their family members as well. It has been well documented in the literature that family and friends play a supportive role for older adults transitioning from one care setting to another and transitioning to living at home (33, 34). It is also well understood that this informal caregiving is stressful and has an impact on the caregivers’ own health and wellbeing (35-38). Although this was not the focus of this study, the value of transitional care interventions delivered by TSOs for family caregivers warrants exploration in future work.

The focus on psychosocial support within the programs included in this study, and the high value placed on this aspect of the program by study participants align with current literature on the importance of social supports (39, 40). Social isolation and loneliness are associated with poor physical and mental health outcomes, including higher rates of depression, and cognitive decline (41). This is a particular risk for older adults after hospital discharge, and evidence suggests that volunteers can provide this social support as part of a multifaceted program for community dwelling older adults (42).

As health systems worldwide tackle social isolation and re-structuring healthcare to improve health and social care integration (43), the voluntary sector may become an invaluable partner that can provide services not necessarily provided by health system partners. Our study findings showed that the services of TSOs tend to focus on psychosocial support and community participation; needs are often of secondary concern compared to improved safety and physical independence once a client has returned home (44). Most health system transitional support services do not address social problems directly, but rather they refer clients to TSOs, such as those included in this study, for activities focused on community engagement and participation (45, 46). Miller & colleagues (2013), in their review of TSO-commissioned preventative care services, noted that the services generally focused on supporting older people from becoming ill, avoiding frailty, reducing the impacts of existing conditions, and supporting a return to independence following an illness; social isolation was a priority focus of these community-based services (47). Rather than expanding existing transitional care programs to address these needs, or leaving these needs unaddressed, this may create an opportunity for greater collaboration with TSO whereby existing programs and professional staff could remain focused on needs related to functional ability and independence in the home. At the same time, TSOs and their volunteers are engaged to provide support with instrumental activities of daily living and facilitating social participation outside the home.

The participation of TSOs in Anglo-American health services has been widely discussed (48-51). The engagement of these agencies within health systems has often been instigated as a response to health system reforms or constrained resources, with health system leaders turning to partnerships to support improved patient care (52) or to expand the breadth and quality of health and social services (53-58)

Although the academic literature focused on TSOs specific to health care transitions is limited, the concept is emerging (59); rather, there has been increasing interest in leveraging volunteers to be engaged in intermediate care (a broad term used to describe a range of transitional care models), particularly in supporting patients transitioning between acute, primary and social care sectors. Furthermore, in 2017, the National Institute for Health and Care Excellence in UK published a guideline on intermediate care models (11) and identified the need for evidence regarding team structure and composition, noting that “the evidence on views and experiences of home-based intermediate care is exclusively from health and social care practitioners, with no evidence from other care and support practitioners from the community” National Insititute for Health and Care Excellence, 2017). Subsequently, in 2020, an international Delphi study sought to define intermediate care, resulting in the following definition: “Intermediate care is a broad range of time-limited services, from crisis response to support for several weeks or months, that aims to ensure continuity and quality of care and promote recovery at the interface between hospital and home, care home, primary care and community services” (60). Importantly the definition also delineated that an interdisciplinary team best delivers intermediate care within an integrated health and social care system that links different providers and levels of care in a collaborative network of care and support that includes partners from community and voluntary sectors.

Volunteers did not provide any personal care supports as these activities may be outside their scope. In the context of strained health system resources, TSOs and their volunteers may be able to provide many of the non-clinical aspects essential to successful transitions from hospital to home, effectively extending the reach of public sector-funded services and providing programs and services most responsive to the unique needs of their community members. Volunteers within TSOs are most often from the communities they serve and thus possess a unique understanding of the needs of their community members as well as community resources (50, 58, 61). Additionally, with this knowledge, they may be well positioned to match older adults to community resources required to live independently and with meaning. Each program included in this study drew volunteers from their respective communities. Regarding time-limited services, they provided referrals or signposting to other locally available supports that may meet clients’ longer-term needs.

Despite the engagement of TSOs, as a sector, they only represent a small portion of service delivery, even in countries with robust voluntary sector engagement in health services (62, 63). It is also important to note that the engagement of TSOs, and the services they provide within their communities, is highly dependent on the broader socio-political environment they are situated within. For example, Grassman (2006) questioned the role of the third sector in Nordic health systems, given their lack of involvement in service delivery to the degree to which TSOs have been in Anglo-American countries (64). Thus, existing literature must be considered within the socio-political context it was generated. This study’s topic is framed within British and Anglo-American conceptualization of third-sector engagement in health service delivery. Broader macro-level discussions of the third sector’s role in providing health services will be undertaken elsewhere.

## Conclusion

In conclusion, this study provides an overview and discussion of four transition support programs delivered by TSOs. Through the analysis and discussion of these types of services, we have highlighted the value of these programs for older adults transitioning from hospital to home and returning to independent living. The findings suggest that the programs can provide patients with a ‘safety net’ and promote independent living. Having skilled volunteers can positively impact older adults’ experience returning home. When the programs under study are considered in tandem with existing evidence, it facilitates a discussion of how TSO services could be made available more widely to support older adults in their transition experiences.

## Data Availability

All data produced in the present study may be available upon reasonable request to the authors

